# Differences in Bladder Cancer Diagnosis by Demographic Factors: A Simulation Modeling Analysis

**DOI:** 10.64898/2026.01.30.26344972

**Authors:** Praveen Kumar, Fernando Alarid-Escudero, Tanvi V Chiddarwar, David Ulises Garibay-Treviño, Krishna Roy Chowdhury, Prince Peprah, Bruce L Jacobs, Karen M Kuntz, Stella K. Kang, Thomas A. Trikalinos, Hawre Jalal

**Affiliations:** Department of Health Policy and Management, School of Public Health, University of Pittsburgh; Public Health Dynamics Laboratory, School of Public Health, University of Pittsburgh; Department of Health Policy, School of Medicine, Stanford University; Division of Health Policy and Management, School of Public Health, University of Minnesota; School of Epidemiology and Public Health, Faculty of Medicine, University of Ottawa; Department of Urology, Division of Health Services Research, University of Pittsburgh; Department of Radiology, Columbia University Irving Medical Center; Department of Health Services, Policy, and Practice and Center for Evidence Synthesis in Health, School of Public Health, Brown University

**Keywords:** Bladder cancer, disparities, model, diagnosis, simulation

## Abstract

**Purpose:** Bladder cancer is associated with significant morbidity and mortality in the US, with 85,000 new cases and 17,400 deaths expected in 2025. Black patients are more likely than White patients to be diagnosed with bladder cancer at advanced stages, as are female patients compared with male patients. We examine whether differences in cancer diagnosis rates by race and sex can explain the observed variability using a simulation model and project outcomes of potential improvement in diagnosis.

**Methods:** We developed a state transition model for bladder cancer to simulate four cohorts based on sex (males, females) and race (Blacks, Whites) from birth through various health states, including disease-free, preclinical stages (0a/0is - IV), clinical stages (0a/0is - IV), and death (bladder cancer or other cause death). Parameters related to disease onset, progression, and diagnosis were estimated by calibrating the model to race- and sex-specific incidence rates by age, and stage distribution at diagnosis for cases diagnosed between 2015 and 2019 in SEER 17 registry areas. We conducted a scenario analysis to examine the impact of differences in diagnosis rates on stage distribution and life expectancy, assuming that Black males (or females) and White females had diagnosis rates similar to those of White males.

**Results:** The calibrated model attributes the differences in stage distribution to lower diagnosis rates in White females (hazard ratio, [HR] = 0.95, 95% credible interval [CI]: 0.92 - 0.96), Black males (0.80, 95% CI: 0.75 - 0.81) and Black females (0.56, 95% CI: 0.53 - 0.58), relative to White males. If diagnosis rates for all demographic groups were similar to White males, the expected life span of a 65-year-old bladder cancer patient would increase by 0.2 years for White females (from 13.8 to 13.9 years), 0.6 years for Black males (from 10.6 to 11.1 years), and 1.9 years for Black females (from 10.5 to 12.4 years).

**Conclusions:** Differences in diagnosis rates of bladder cancer by race and sex explain the observed differences in stage distribution at diagnosis. Targeted interventions aimed at improving diagnosis rates have the potential to substantially improve survival for patients with bladder cancer.

## INTRODUCTION

In 2025, an estimated 85,000 new bladder cancer cases and 17,400 deaths are expected in the United States.^1^ The disease is about twice as common in White males as in Black males.^2^ However, Black patients are more likely to be diagnosed at advanced stages^3–7^ and to experience poorer survival, even after controlling for stage, compared with White patients.^6,8^ These disparities in diagnosis are particularly pronounced among Black females, where 36% are diagnosed with muscle-invasive bladder cancer compared with 24% for White females.^9^

The possible explanations for delayed diagnosis among females include misattribution of symptoms to urinary infections.^10–13^ In contrast, advanced-stage diagnosis among Black patients is likely driven by barriers to accessing care, a higher risk of developing aggressive forms of bladder cancer (e.g., squamous cell and adenocarcinomas), and a greater likelihood of high-grade disease.^4,5^ Although several studies have consistently documented diagnostic delays,^3–5^ no prior study has estimated differences in diagnosis rates by race and sex. Such information is critical for understanding the magnitude of differences and guiding targeted interventions to reduce diagnostic delays.

Simulation models can be used to estimate differences in diagnosis rates by evaluating plausible scenarios and identifying those that best reproduce observed data on age-specific incidence and stage distribution across population subgroups (White males, White females, Black males, and Black females). These models are particularly valuable for problems in which key biological processes – such as cancer onset, progression, and diagnosis – are unobservable, and only outcomes like incidence and stage at diagnosis are observed. By calibrating simulated outcomes to epidemiologic data, these models can reveal differences in rates of disease onset, progression, and diagnosis. Our objective was to use a simulation model to estimate differences in bladder cancer diagnosis rates by race and sex and to estimate the potential for improvement in survival from earlier diagnosis among Black patients and females.

## METHODS

We developed a discrete-time Markov model to simulate the life trajectory of a cohort, producing age-specific incidence rates (40-44, 45-49, …, 85+) and stage distribution (stage 0a/0is, stages I-IV) for newly diagnosed bladder cancer. The model included unobservable parameters related to bladder cancer onset, progression, and diagnosis, which were estimated by calibrating the model against observed incidence rates and stage distributions of diagnosed bladder cancer cases from 2015 to 2019, using the Surveillance, Epidemiology, and End Results (SEER) Program data. We compared the diagnosis rates across four subgroups of interest – White males, White females, Black males, and Black females – to examine differences by race and sex. Finally, to demonstrate the effect of making diagnosis rates similar across all four subgroups, we simulated the expected change in survival if Black populations and females had diagnosis rates comparable to those of White males.

### Model Overview

Our model comprised the following health states: disease-free, five preclinical stages (preclinical stage 0a/0is, preclinical stages I-IV), five clinical stages (stage 0a/0is, stage I-IV), and death (bladder cancer-specific or other cause) (Figure 1). The preclinical stages refer to undiagnosed bladder cancer, while the clinical stages refer to diagnosed cases. We ran the model separately for four cohorts, i.e., White males, White females, Black males, and Black females. The disease process was divided into four modules: onset, progression, diagnosis, and survival.

- Disease onset – The onset rate was modeled as a function of sex, race, and age. Specifically, we used a general phase-type distribution to model the transition from the ‘Disease-free’ state to the ‘Preclinical Stage 0a/0is’ state in two sequential phases.^14^ The first phase (from disease-free to an intermediate state) assumed a constant hazard, and the second phase (from intermediate to the preclinical stage 0a/0is state) used a Weibull hazard. This structure increased model flexibility, enabling us to better capture age-dependent risk of disease onset by sex and race, and improving overall model fit. Further details on the method are provided in the Supplementary Appendix.
- Progression – In the preclinical stage 0a/0is, bladder cancer can progress further to subsequent preclinical stages I-IV. We modeled such transitions to occur at constant hazard rates, which were assumed to be the same across the four cohorts, because there is no evidence suggesting that bladder cancer progresses differently by race and sex.
- Diagnosis – Bladder cancer can be diagnosed at any preclinical stage. Because the likelihood of diagnosis increases with advancing stage, we modeled stage-specific diagnosis rates that were constrained to increase with stage. In addition, these rates were allowed to vary by sex and race.
- Survival – People can die from two causes: bladder cancer and other causes. For bladder cancer, we used cause-specific death rates stratified by stage, sex, and race of the cohort, obtained from the SEER registry data. For other-cause mortality, we used sex- and race-specific US life tables from 2017.^15^

**Figure 1.**
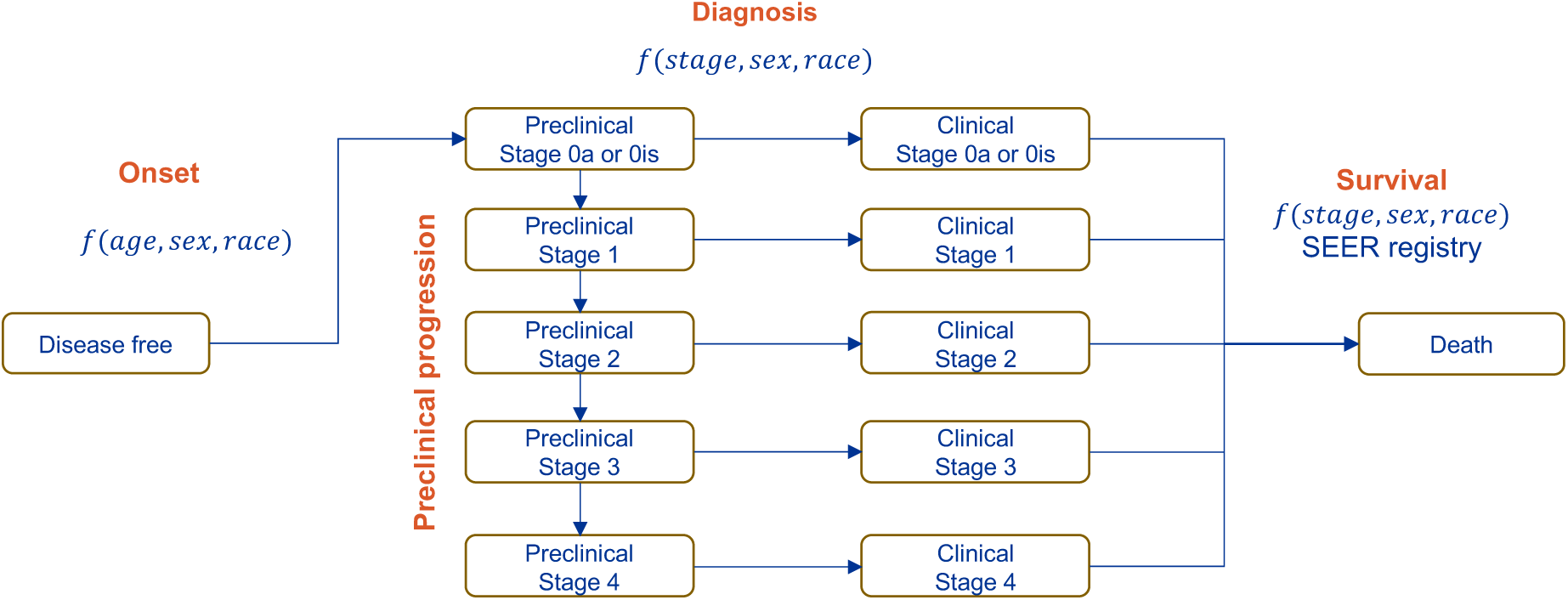
Schematic of the cohort simulation model. The model comprises 12 health states – disease-free, five preclinical stages, five clinical stages, and death – represented as boxes. Arrows indicate the permissible transitions between states. The model includes four submodules: disease onset, preclinical progression, diagnosis, and survival. Onset rates vary by age, sex, and race. Diagnosis rates depend on stage, sex, and race, with more advanced stages having a higher likelihood of diagnosis. Survival also varies by stage, sex, and race.

### Data

The SEER Program is a surveillance system coordinated by the National Cancer Institute that routinely collects information on newly diagnosed cancer cases residing in SEER registry areas.^9^ Using SEER 17 registry data, we sourced i) incidence rate of bladder cancer by age group (40-44, 45-49, …, 80-84, 85+), sex (males, females), race (Black, White), ii) sex and race-specific stage distribution at diagnosis, and iii) cause-specific survival of bladder cancer by stage, sex and race. The incidence rates were used as calibration targets to estimate unobservable parameters related to disease onset, progression, and diagnosis, as described in the *Calibration* section.

We chose SEER Program’s specialized database^9^, SEER Research Plus Data (Specialized Staging Over Time Fields) because it accounts for changes in disease definition over time,^16^ allowing incidence data to be pooled across multiple years. A bladder cancer case was identified using the following criteria: Primary Site codes (C67.0-C67.9), Behavior recode for analysis (‘Malignant’), and ICD-O-3 histology codes specific to transitional cell carcinoma (8120, 8122, 8130). We focused on cases diagnosed between 2015 and 2019 and excluded cases diagnosed at autopsy or through death certificates.

### Calibration

The model (M) included 20 unobservable parameters – onset (8 parameters), progression (4 parameters), and diagnosis (8 parameters) (Table 1). We estimated these parameters using a Bayesian approach to obtain their joint posterior distribution. Specifically, we applied the incremental mixture importance sampling (IMIS) method,^17,18^ which efficiently samples from regions of the parameter space with higher posterior density. Non-informative priors were specified as uniform distributions. We defined the likelihood function assuming that each calibration target *t* followed a normal distribution with mean *Q_t_* and standard deviation σ*_t_*.

**Table 1.** Comparison of lifetime risk estimates from the model and from DevCAN.

| **Cohort** | **Model-based<sup>1</sup>** | **DevCAN<sup>2</sup>** |
| --- | --- | --- |
| White males | 3.75% (3.73%, 3.79%) | 3.85% |
| White females | 1.06% (1.04%, 1.07%) | 1.18% |
| Black males | 1.63% (1.59%, 1.68%) | 1.72% |
| Black females | 0.66% (0.63%, 0.68%) | 0.75% |

<sup>1</sup> The values in parentheses represent the 95% credible interval
<sup>2</sup> DevCAN estimates are based on bladder cancer cases diagnosed in 2017-19.

The mean *Q_t_* refers to the expected value of the model-predicted output for a parameter set θ, i.e., *E*[*M*(θ)]. Table S1 lists all the parameters estimated by calibration along with their prior distributions. We evaluated the performance of the calibration by i) comparing the model output of incidence rates and stage distribution against the actual data by sex and race, ii) reviewing the posterior distribution against the prior distribution, and iii) analyzing the pairwise correlation among calibrated parameters in the posterior distribution.

For validation, we compared the model-predicted lifetime risk across the four subgroups with estimates from DevCan^19^ software from the National Cancer Institute, which reports the lifetime risk of bladder cancer diagnosis by years of diagnosis.

### Scenario analysis

We ran hypothetical scenarios to examine the impact of differences in diagnosis rates on stage distribution, assuming that Black males (or females) and White females had diagnosis rates similar to those of White males (the reference group). These scenarios were based on 500 sampled parameter sets from the joint posterior distribution of calibrated parameters. We then calculated the expected change in life expectancy resulting from the shift in stage distribution. For example, if diagnosis rates among Black males were equal to those of White males, a higher proportion of cases would be diagnosed at earlier stages compared to the actual data. This shift in stage distribution would likely result in a survival benefit, as lower stages are associated with better survival outcomes. To estimate the change in survival, we used race-, sex-, and stage-specific survival rates.

## RESULTS

### Model Fit

Figure 2A shows the model-predicted versus observed age-specific incidence rates for all bladder cancers diagnosed among White males, White females, Black males, and Black females during 2015–2019. Similarly, Figure 2B presents the model-predicted versus observed stage-specific incidence for the same groups over the same period. For nearly all age groups and across all models, the uncertainty intervals for the predicted and observed incidences overlap, indicating that the model closely reproduces the epidemiologic data.

**Figure 2.**
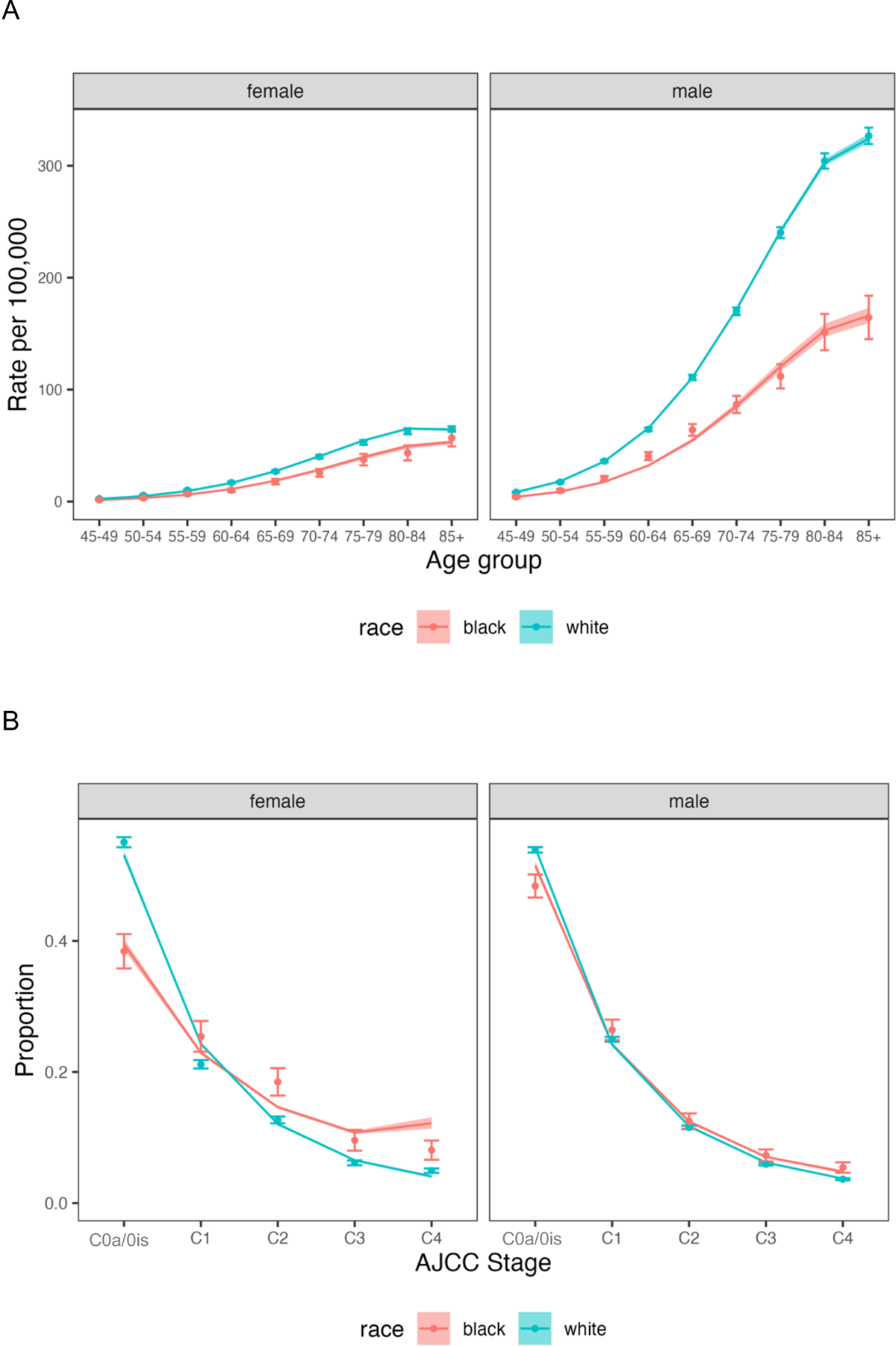
Model Fit to Observed Data. The figure presents the comparison between the model-predicted and observed bladder cancer incidence and stage distribution. Predicted values are shown as colored lines with shaded areas indicating the 95% credible intervals. Observed data are shown as dots with 95% confidence intervals (error bars): (A) age-specific incidence rates and (B) stage distribution at diagnosis.

Furthermore, the substantially narrower posterior distributions relative to the priors indicate that these parameters are primarily informed by the calibration targets (Figure S2).

### Model Validation

Table 1 compares the model-estimated lifetime risk of being diagnosed with bladder cancer with corresponding estimates from DevCAN. Overall, the two sets of estimates are similar, although the model consistently predicts slightly lower risks than DevCAN. The model-predicted lifetime risks are 3.8% (vs 3.9%) for White males, 1.1% (vs 1.2%) for White females, 1.6% (vs 1.7%) for Black males, and 0.7% (vs 0.8%) for Black females.

### Differences in diagnosis rates by race and sex

Figure 3 presents the hazard ratios (HRs) for bladder cancer diagnosis across population subgroups, using White males as the reference category. For a given stage, White females had a HR of 0.95 (95% credible interval: 0.92 - 0.96), indicating a 5% lower rate of diagnosis compared with White males. In contrast, Black males and Black females showed substantially lower rates of diagnosis, with HRs of 0.80 (95% credible interval, CI: 0.75 - 0.81) and 0.56 (95% CI: 0.53 - 0.58), respectively. Figure S1 displays the distribution of these hazard ratios along with the posterior distributions of other calibrated parameters.

**Figure 3.**
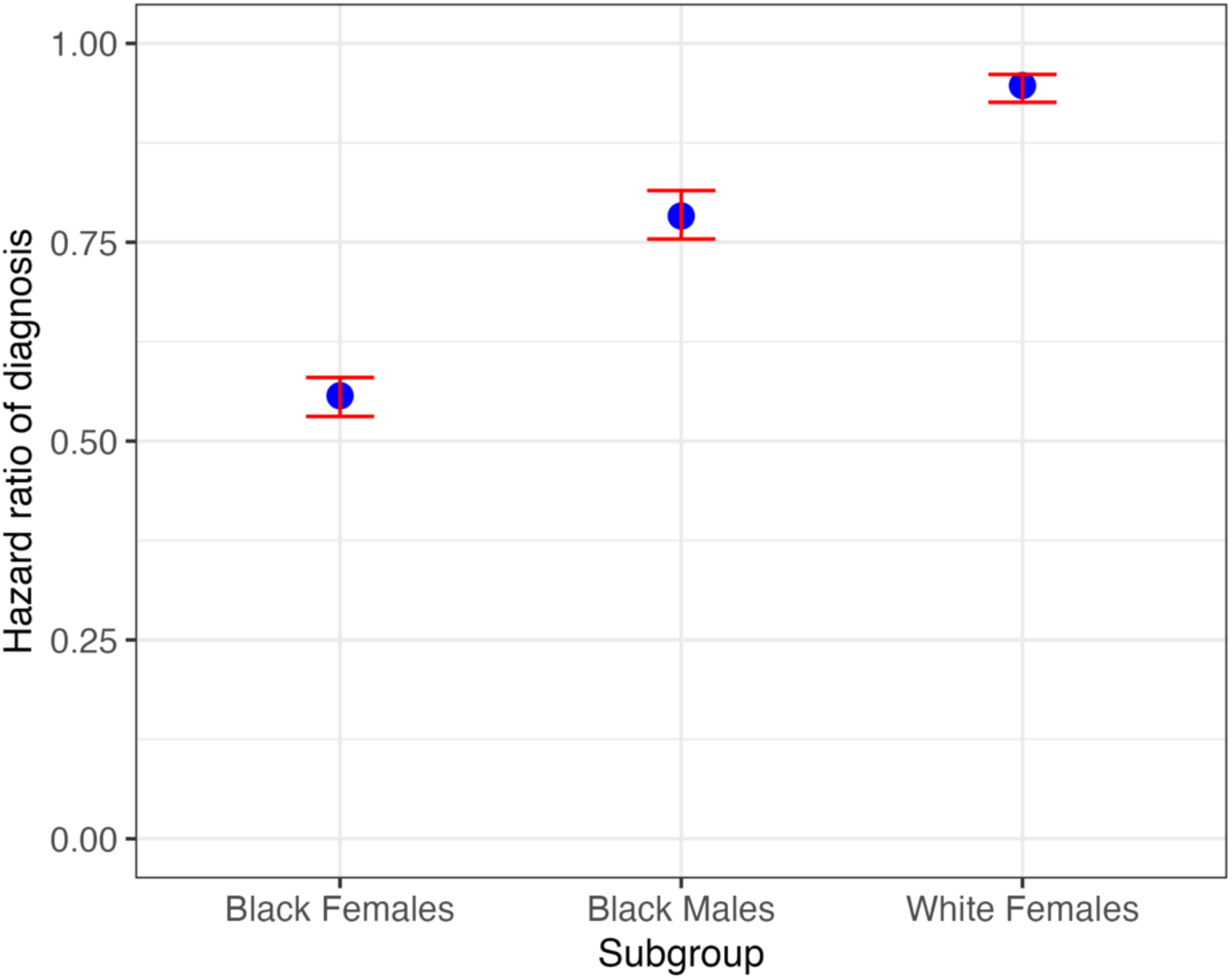
Hazard ratio of diagnosis by subgroups. The plot displays hazard ratios for diagnosis, with White males serving as the reference group. A value of <1 denotes a lower diagnosis rate compared with White males at the same stage of bladder cancer. Mean estimates are shown as blue dots, with 95% credible intervals shown as red bars.

### Probability of diagnosis

Figure S3 shows the probability of diagnosis by age, stage, sex, and race. Specifically, it depicts the annual probability of transitioning from a preclinical stage to the corresponding clinical stage. Across all subgroups, the probability of diagnosis increases with disease stage. For example, a 70-year-old White male patient with preclinical stage 2 bladder cancer has an estimated 30% probability of diagnosis within the next year, which rises to nearly 50% for preclinical stage 4. The probabilities vary by race and sex: White females, Black males, and Black females have lower probabilities of diagnosis, consistent with the patterns shown in Figure 2. Finally, diagnosis probabilities decline with advancing age due to competing risks from non-bladder cancer mortality, a pattern that becomes especially pronounced after age 80 when competing mortality increases substantially.

### Counterfactual analysis

Figure 4 illustrates how equalizing diagnosis rates to those of White males affects bladder cancer mortality across other demographic groups. Under this scenario, mortality risk for a 65-year old bladder cancer patient is projected to decrease by an average of 3% for White females, 15% for Black males, and 41% for Black females. These reductions translate into improved survival outcomes for bladder cancer patients. Table 3 presents the corresponding changes in expected life span when diagnosis rates are set equal to those of White males. As shown in Table 2, the model predicted expected life span of a 65-year-old bladder cancer patient would increase by 0.17 years (95% CI: 0.13 – 0.23) for White females (from 13.8 to 13.9 years), 0.6 years (95% CI: 0.5 – 0.7) for Black males (from 10.6 to 11.1 years), and 1.9 years (95% CI: 1.7 – 2.0) for Black females (from 10.5 to 12.4 years).

**Figure 4.**
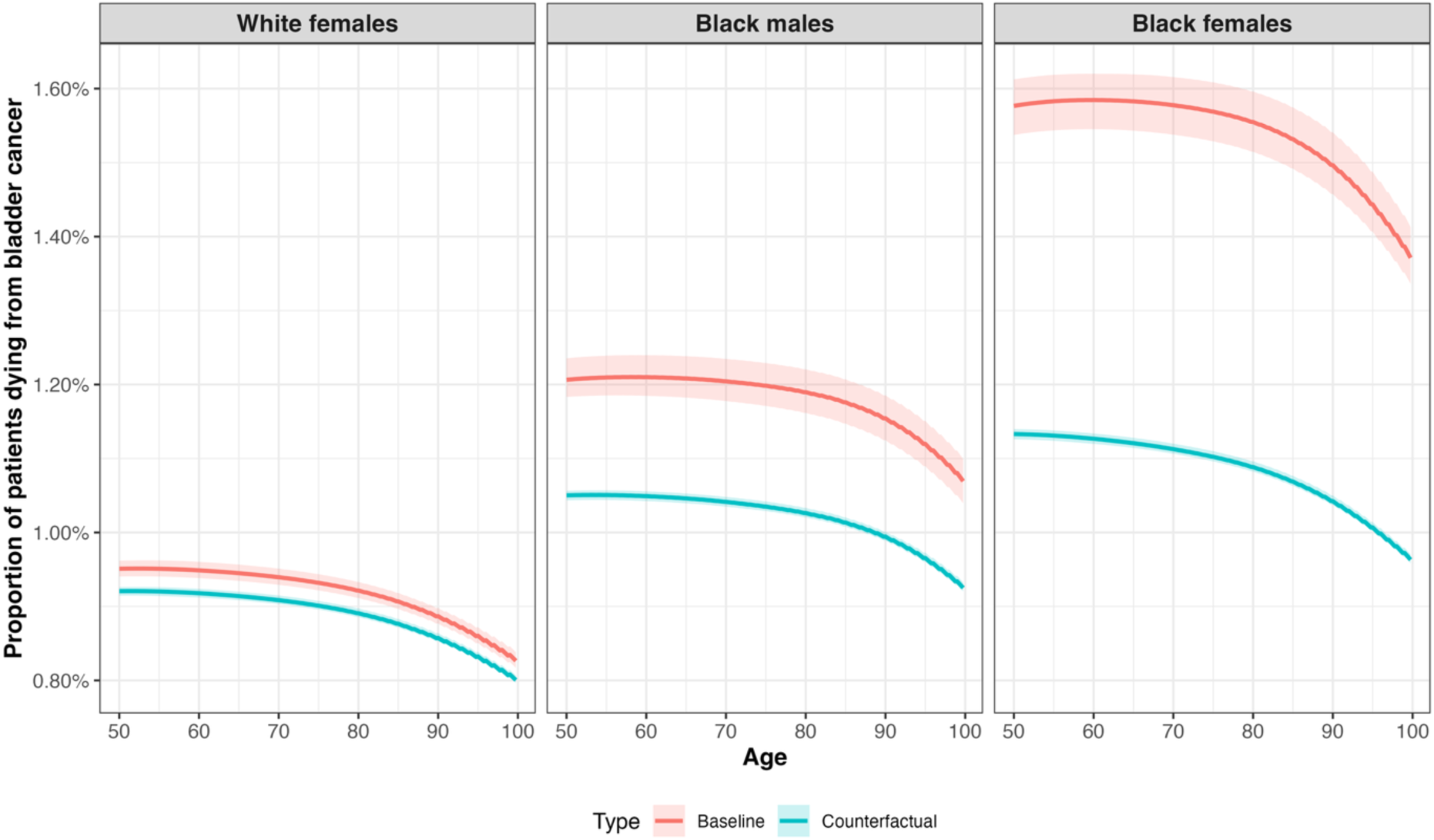
Results of the counterfactual analysis showing bladder cancer mortality if their diagnosis rates were similar to White males. Each plot displays the model-predicted 3-month bladder cancer mortality risk by age for different demographic subgroups under the assumption that all subgroups share the same diagnosis rate as White males. The baseline (red line) refers to the model-predicted mortality risk using the race- and sex-specific diagnosis rates, while the counterfactual (blue line) refers to the model-predicted mortality risk when the diagnosis rate is set equal to that of White males. Shaded areas around the lines indicate the 95% credible intervals.

**Table 2.** Counterfactual Analysis Results: Expected Life Span as Outcome.

| Cohort | Bladder Cancer Patient Age = 65 years<sup>1</sup> |  |  |
| --- | --- | --- | --- |
|  | Baseline | Counterfactual | Difference |
| White Females | 13.8 (13.7, 13.8) | 13.9 (13.9, 14.0) | 0.17 (0.13, 0.23) |
| Black Males | 10.6 (10.5, 10.7) | 11.1 (11.1, 11.2) | 0.6 (0.5, 0.7) |
| Black Females | 10.5 (10.4, 10.7) | 12.4 (12.37, 12.43) | 1.9 (1.7, 2.0) |
| Cohort | Bladder Cancer Patient Age = 75 years<sup>1</sup> |  |  |
|  | Baseline | Counterfactual | Difference |
| White Females | 9.9 (9.8, 9.9) | 10.0 (9.9, 10.0) | 0.08 (0.06, 0.12) |
| Black Males | 7.9 (7.8, 7.9) | 8.2 (8.1, 8.2) | 0.3 (0.2, 0.4) |
| Black Females | 8.2 (8.1, 8.3) | 9.3 (9.2, 9.3) | 1.1 (0.9, 1.1) |

<sup>1</sup> Estimates of expected life span in baseline and counterfactual scenarios are model-based but differ in diagnosis rates. ‘Baseline’ refers to model-predicted life span using race- and sex-specific diagnosis rates, while ‘counterfactual’ refers to predicted life span assuming diagnosis rates equal those of White males. We calculated these outcomes for two age groups: bladder cancer patients at ages 65 and 75. The values represent the mean estimates, with 95% credible intervals shown in parentheses.

## DISCUSSION

Black and female patients are diagnosed at more advanced stages of bladder cancer than White males. We used a simulation model to examine differences in diagnosis rates of bladder cancer by race and sex. The model reflects our conceptual understanding of the data-generating process and allows us to estimate unobservable disease onset, progression, and diagnosis rates by calibrating to observed age-specific incidence and stage distribution at diagnosis. Our main finding is that diagnosis rates differ substantially by race and sex: hazard ratios are 0.95 for White females, 0.80 for Black males, and 0.55 for Black females, relative to White males.

The simulation-based approach enables comparisons across demographic groups that would be infeasible in clinical or empirical studies (due to cost and ethical concerns), as such data would require a longitudinal cohort of healthy individuals followed through disease onset, preclinical progression, and diagnosis. Similar modeling methods have been used in other cancers; for example, Huber (2023) used a similar approach to examine whether race and sex-based disparities in the incidence of multiple myeloma (MM) are attributable to differences in MGUS incidence (a precursor state to MM) or to increased progression rates from MGUS to MM.^20^ Batina (2013), using a microsimulation model, found that breast cancer tumors in Black patients are more aggressive than those in White patients, with faster growth and earlier metastasis.^21^ Mandelblatt (2023) found that treatment related factors contributed most to racial disparities in breast cancer mortality across three independent population models.^22^

Our results indicate that Black females have the lowest diagnosis rate, suggesting they experience the greatest disadvantage. Prior studies note two contributing factors. First, women are often diagnosed at later stages of bladder cancer because their symptoms are frequently misattributed to urinary tract infections.^10–12^ Second, Black patients, on average, face later diagnoses, which could be due to reduced access to timely care, socioeconomic factors, lack of education about signs and symptoms, and more aggressive disease.^4,5^ Together, these factors place Black females at the highest risk of delayed diagnosis. Similar findings have been reported for breast cancer, where Black females experienced the most delays in initial diagnosis for breast cancer, compared to White females.^23–25^

Reducing these differences could substantially improve survival among bladder cancer patients, particularly Black males and females. We estimate a potential survival difference of 1.9 years for Black females and 0.6 years for Black males if their diagnosis rates were equivalent to those of White males. These gains are considerable when compared with improvements achieved through other interventions, such as adjuvant chemotherapy for node-negative breast cancer (life year gain: 0.6-0.9 years),^26^ treatment of hypertension in 40-year old patients (life year gain of 0.75 in females and 1.7 years in males),^27^ and appendectomy in patients with probable acute appendicitis (life year gain: 0.75-2.6 years).^26^ Earlier-stage detection would also reduce the number of cases diagnosed at stages III and IV, which are associated with greater morbidity and higher treatment costs.

However, our analysis does not identify the underlying mechanism driving these disparities in diagnosis rates. These differences are likely influenced by a combination of system-level (e.g., access to care, insurance coverage), patient level (health seeking behavior, natural history of disease, health literacy) and environmental factors (e.g., exposure to risk factors). Identifying the pathways that contribute to these differences is an important area for future research and may inform the development of targeted interventions. Examples of potential strategies include: considering bladder cancer as a potential diagnosis in women and Black patients referred to oncology with hematuria; implementing health literacy communication and bladder cancer awareness campaigns in communities with high proportions of Black residents; and building relationships with primary care, OB/GYN and urology providers to improve access to care and reduce delays in referral.^28^

Our study has several limitations. First, Black populations are disproportionately affected by more aggressive and less common bladder cancer histologies, including squamous cell carcinoma (SCC) and adenocarcinoma.^4,29^ Because these subtypes are rare – SCC accounts for 2-5% of bladder cancers and adenocarcinoma for <2%^4,29,30^ – our analysis was restricted to TCC, which represents approximately 93% of cases. As a result, our findings may not fully capture differences in disease biology or progression patterns associated with non-TCC histologies. We therefore implicitly assume similar preclinical progression rates for TCC across demographic subgroups. It is theoretically possible that yet unidentified factors associated with sex and race explain at least part of the observed differences. However, there are no clear candidates for such factors, and, as shown in this work, they are not necessary to explain the observed differences in stage at diagnosis by sex and race.

Second, we did not include tumor grade in the model. Since prior studies have reported a higher proportion of high-grade tumors among Black patients compared with White patients,^5,31^ we are interested in examining whether including grade implies that the observed differences by sex and race can no longer be explained by differences in diagnosis rates. We will pursue this analysis in a comparative modeling framework.^32^ Third, we did not explicitly model the effect of risk factors such as smoking or other carcinogens, which differ across demographic subgroups. Instead, we used subgroup-specific onset functions, thereby implicitly capturing variation in exposure levels. This approach limits our ability to directly evaluate the role of subgroup-specific risk factor distributions and represents an important area for future work.

## CONCLUSION

We found variation in diagnosis rates that can explain the higher burden of bladder cancer among Blacks and women, and addressing these delays can help achieve substantial gains in life expectancy among these patients. Targeted efforts to understand the mechanism and implement interventions to reduce these differences are essential to improving clinical outcomes for affected populations.

## Data Availability

All data produced in the present study are available upon reasonable request to the authors. The data used in this study are available in the SEER database, however the harmonized staging variables, also called as staging over time variables, are available by request to the SEER Program team. More details about the data and how to access it can be found at https://seer.cancer.gov/data/specialized/available-databases/staging-time-request/.

## Acknowledgments

This study is supported by the grant U01CA265750 from the National Cancer Institute (NCI) as part of the Cancer Intervention and Surveillance Modeling Network (CISNET). The content is solely the responsibility of the authors and does not necessarily represent the official views of the National Institutes of Health. The funding agency had no role in the study’s design, interpretation of results, or writing of the manuscript. The funding agreement ensured the authors’ independence in designing the study, interpreting the data, writing, and publishing the report. The author acknowledges the use of ChatGPT, a generative AI tool, to assist in refining sentence structure, improving clarity, and ensuring conciseness in the manuscript. All intellectual content, critical analysis, and interpretations remain the sole work of the author.

## Supplementary Appendix

### Bladder cancer onset

To model bladder cancer onset, we used a general phase type (PH) distribution with two steps. This provides flexibility in shaping the hazard of disease onset by incorporating additional parameters governing the transition from the disease-free state to the preclinical stage 0a/0is. Specifically, the time to onset of the preclinical cancer becomes a convolution of two random processes, i.e., from i) disease-free to intermediate state, and ii) intermediate to preclinical stage 0a/0is. The transition from a disease-free to an intermediate state is assumed to occur at constant transition rates that vary by race and sex of the cohort. However, the transition from intermediate to preclinical stage is assumed to depend on sex and age.

---

**Figure S1:**
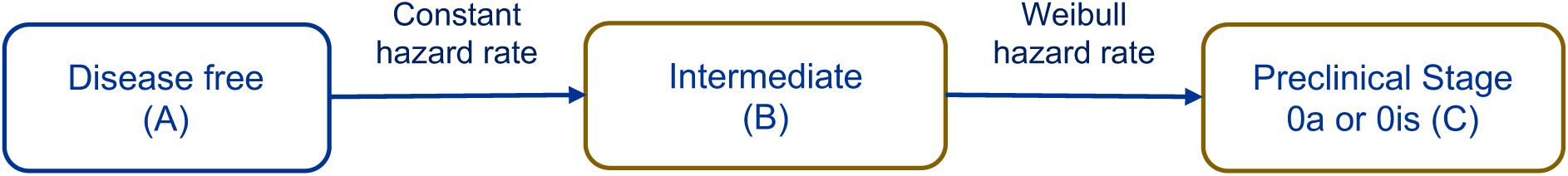
Disease onset module

**Figure S2.**
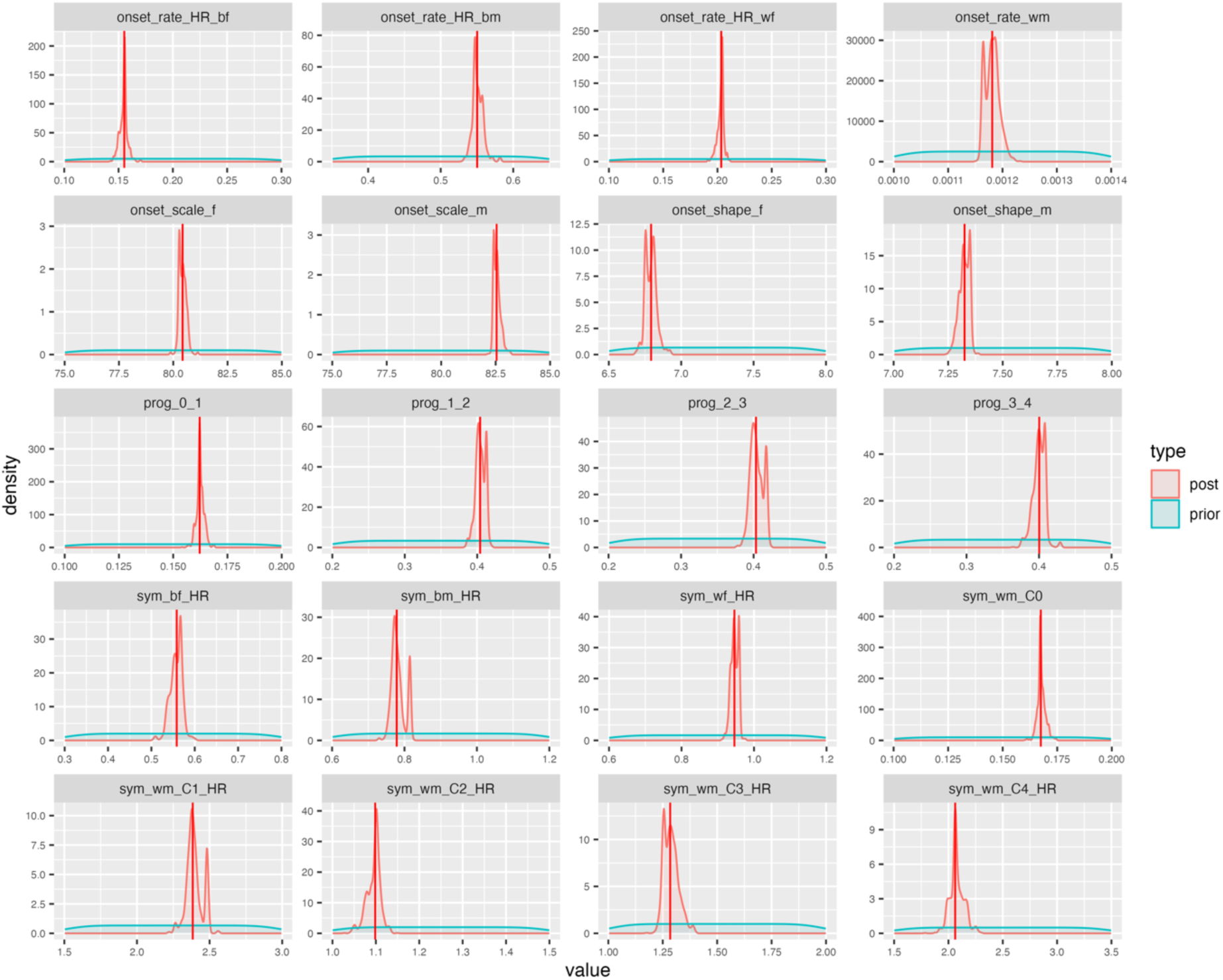
Posterior distribution of calibrated parameters.

**Figure S3.**
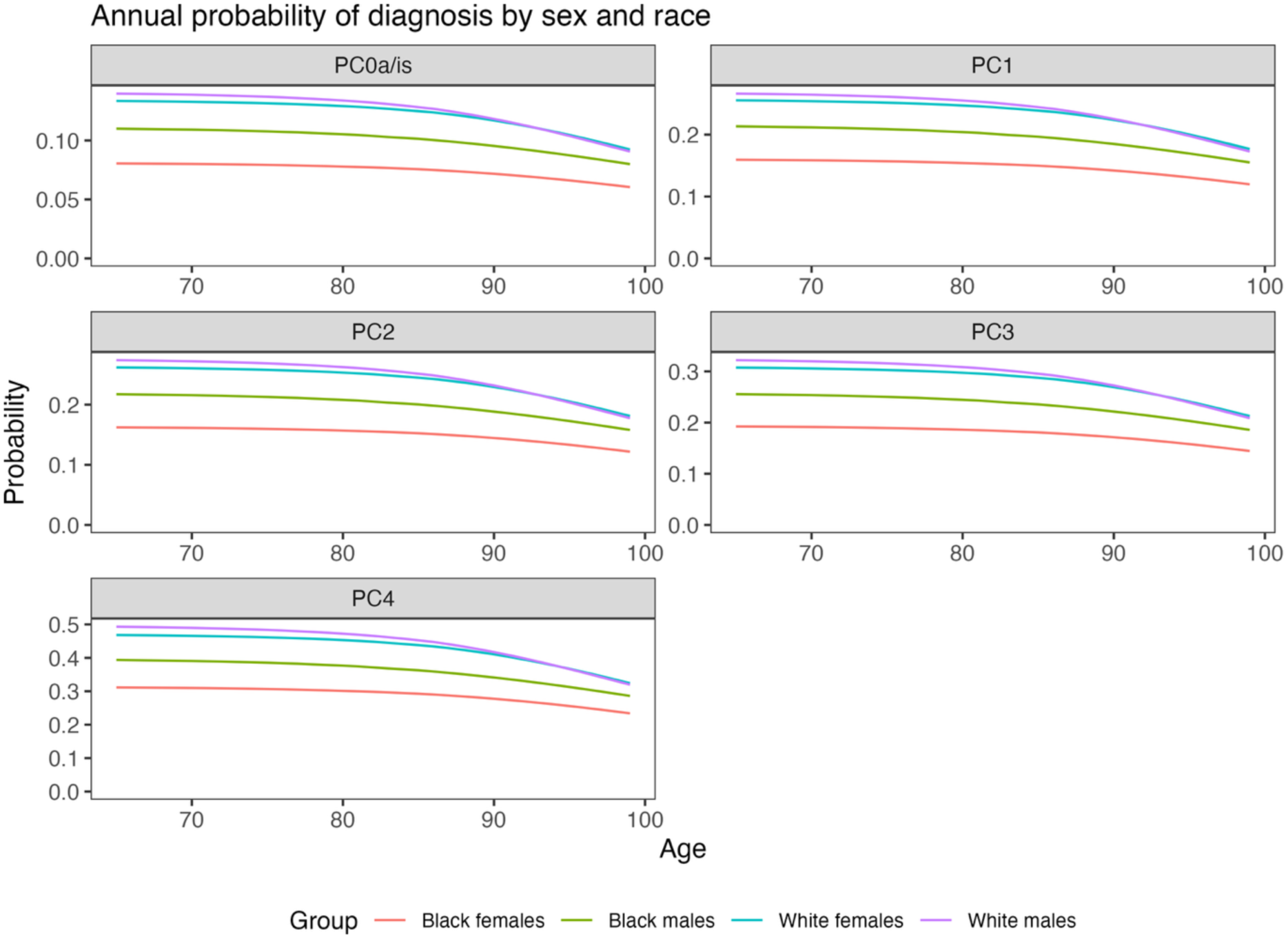
Age-specific probability of diagnosis by sex, and race. The plots show the annual probability of transitioning from preclinical stage *i* to the corresponding clinical stage *i*. Within each plot, different-colored lines represent demographic subgroups: White males (purple), White females (blue), Black males (green), and Black females (red).

**Table S1.**
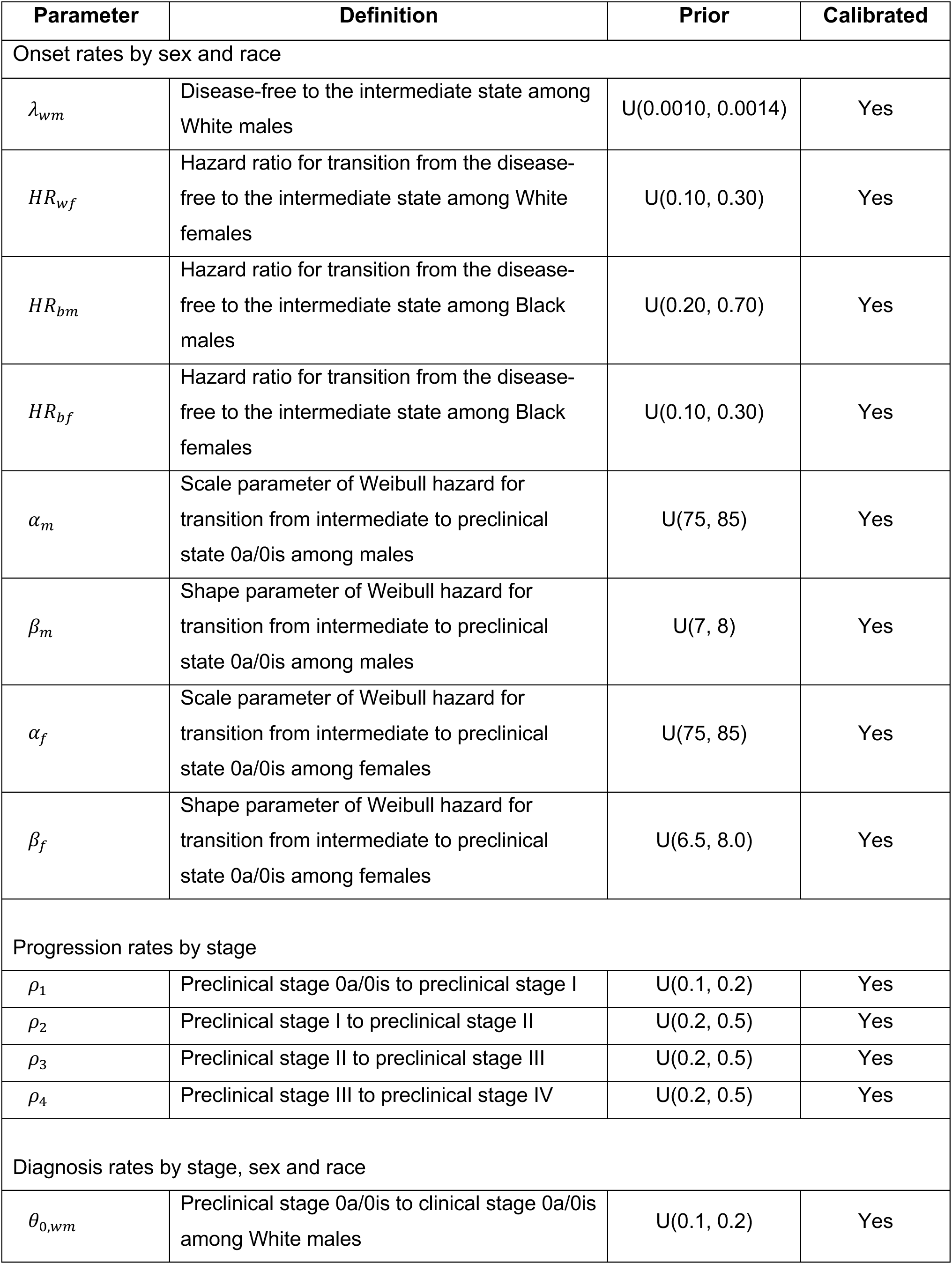
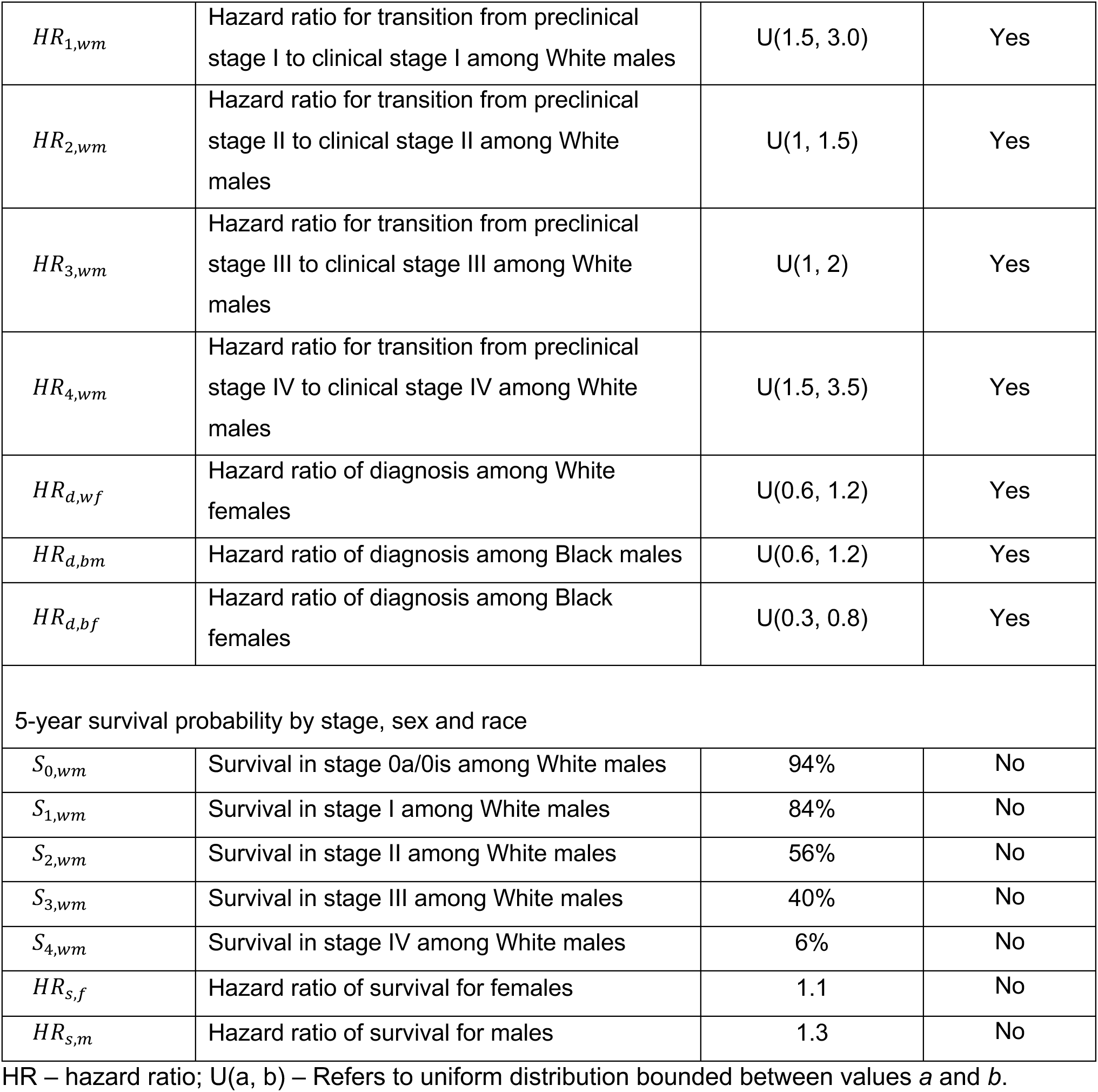
Parameter definitions and prior distributions.

| Parameter | Definition | Prior | Calibrated |
| --- | --- | --- | --- |
| Onset rates by sex and race |  |  |  |
| $\lambda_{wm}$ | Disease-free to the intermediate state among White males | U(0.0010, 0.0014) | Yes |
| $HR_{wf}$ | Hazard ratio for transition from the disease-free to the intermediate state among White females | U(0.10, 0.30) | Yes |
| $HR_{bm}$ | Hazard ratio for transition from the disease-free to the intermediate state among Black males | U(0.20, 0.70) | Yes |
| $HR_{bf}$ | Hazard ratio for transition from the disease-free to the intermediate state among Black females | U(0.10, 0.30) | Yes |
| $\alpha_m$ | Scale parameter of Weibull hazard for transition from intermediate to preclinical state 0a/0is among males | U(75, 85) | Yes |
| $\beta_m$ | Shape parameter of Weibull hazard for transition from intermediate to preclinical state 0a/0is among males | U(7, 8) | Yes |
| $\alpha_f$ | Scale parameter of Weibull hazard for transition from intermediate to preclinical state 0a/0is among females | U(75, 85) | Yes |
| $\beta_f$ | Shape parameter of Weibull hazard for transition from intermediate to preclinical state 0a/0is among females | U(6.5, 8.0) | Yes |
| Progression rates by stage |  |  |  |
| $\rho_1$ | Preclinical stage 0a/0is to preclinical stage I | U(0.1, 0.2) | Yes |
| $\rho_2$ | Preclinical stage I to preclinical stage II | U(0.2, 0.5) | Yes |
| $\rho_3$ | Preclinical stage II to preclinical stage III | U(0.2, 0.5) | Yes |
| $\rho_4$ | Preclinical stage III to preclinical stage IV | U(0.2, 0.5) | Yes |
| Diagnosis rates by stage, sex and race |  |  |  |
| $\theta_{0,wm}$ | Preclinical stage 0a/0is to clinical stage 0a/0is among White males | U(0.1, 0.2) | Yes |
| $HR_{1,wm}$ | Hazard ratio for transition from preclinical stage I to clinical stage I among White males | U(1.5, 3.0) | Yes |
| $HR_{2,wm}$ | Hazard ratio for transition from preclinical stage II to clinical stage II among White males | U(1, 1.5) | Yes |
| $HR_{3,wm}$ | Hazard ratio for transition from preclinical stage III to clinical stage III among White males | U(1, 2) | Yes |
| $HR_{4,wm}$ | Hazard ratio for transition from preclinical stage IV to clinical stage IV among White males | U(1.5, 3.5) | Yes |
| $HR_{d,wf}$ | Hazard ratio of diagnosis among White females | U(0.6, 1.2) | Yes |
| $HR_{d,bm}$ | Hazard ratio of diagnosis among Black males | U(0.6, 1.2) | Yes |
| $HR_{d,bf}$ | Hazard ratio of diagnosis among Black females | U(0.3, 0.8) | Yes |
| 5-year survival probability by stage, sex and race |  |  |  |
| $S_{0,wm}$ | Survival in stage 0a/0is among White males | 94% | No |
| $S_{1,wm}$ | Survival in stage I among White males | 84% | No |
| $S_{2,wm}$ | Survival in stage II among White males | 56% | No |
| $S_{3,wm}$ | Survival in stage III among White males | 40% | No |
| $S_{4,wm}$ | Survival in stage IV among White males | 6% | No |
| $HR_{s,f}$ | Hazard ratio of survival for females | 1.1 | No |
| $HR_{s,m}$ | Hazard ratio of survival for males | 1.3 | No |

HR – hazard ratio; U(a, b) – Refers to uniform distribution bounded between values *a* and *b*.

